# Comparing “people-like-me” and linear mixed model predictions of functional recovery following knee arthroplasty

**DOI:** 10.1101/2022.03.09.22271922

**Authors:** Jeremy Graber, Andrew Kittelson, Elizabeth Juarez-Colunga, Xin Jin, Michael Bade, Jennifer Stevens-Lapsley

## Abstract

**Objective:** Prediction models can be useful tools for monitoring patient status and personalizing treatment in health care. The goal of this study was to compare the relative strengths and weaknesses of two different approaches for predicting functional recovery after knee arthroplasty: a neighbors-based “people-like-me” (PLM) approach and a linear mixed model (LMM) approach.

**Materials and Methods:** We used two distinct datasets to train and then test PLM and LMM prediction approaches for functional recovery following knee arthroplasty. We used Timed Up and Go (TUG)—a commonly used test of mobility—to operationalize physical function. Both approaches used patient characteristics and baseline postoperative TUG values to predict TUG recovery from days 1-425 following surgery. We compared the accuracy and precision of PLM and LMM predictions in the testing dataset.

**Results:** A total of 317 patient records with 1379 TUG observations were used to train PLM and LMM approaches, and 456 patient records with 1244 TUG observations were used to test the predictions. The approaches performed similarly in terms of mean squared error and bias, but the PLM approach provided more accurate and precise estimates of prediction uncertainty.

**Discussion and Conclusion:** Overall, the PLM approach more accurately and precisely predicted TUG recovery following knee arthroplasty. These results suggest PLM predictions may be more clinically useful for monitoring recovery and personalizing care following knee arthroplasty. However, clinicians and organizations seeking to use predictions in practice should consider additional factors (e.g., resource requirements) when selecting a prediction approach.

## BACKGROUND AND SIGNIFICANCE

Prediction models can be useful tools for supporting decision making in healthcare. They can be used to inform patients about the future course of their condition, estimate prognosis, and select and evaluate treatments.^1,2^ Prediction models can account for prognostic differences across individuals and subgroups in a population by adjusting for relevant factors.^2,3^ In doing so, prediction models can facilitate a personalized medicine approach, where treatments and decisions are anchored to an individual’s prognosis instead of the average outcomes of the population.^4^

Prediction models may be particularly useful for personalizing care in postoperative settings where (1) clinical status changes rapidly, (2) early care decisions may have long-term impacts on recovery, and (3) personal factors influence the expected course of recovery. The total knee arthroplasty (TKA) population is a good example to illustrate the usefulness of prediction models in postoperative care because (1) patients experience dramatic functional loss acutely after TKA followed by a period of rapid recovery,^5^ (2) targeted interventions during the acute phase can improve long-term function,^6-9^ and (3) a combination of personal factors (e.g., BMI, sex) can affect an individual’s functional recovery trajectory.^10^ Therefore, prediction models can be used to augment clinical decision making and facilitate personalized treatments to improve functional recovery following TKA.^11^

We recently developed a “people-like-me” (PLM) approach for predicting functional recovery following TKA.^12,13^ We used Timed Up and Go (TUG)—a common clinical test of mobility—to operationalize physical function. In this previous work, our PLM approach used a semi-parametric, neighbors-based prediction method, where a new patient’s TUG recovery was predicted using the observed TUG recovery data of similar historical patients.^14^ We hypothesized this neighbors-based method would facilitate highly realistic and flexible predictions,^15^ which might confer superior accuracy and precision compared to alternative prediction approaches. However, the PLM approach was also computationally intensive and relied on a large donor dataset, which required substantial resources to collect and maintain. Therefore, the purpose of this study was to compare the PLM approach to an alternative, less resource-intensive approach—prediction based on linear mixed models (LMM).^16-18^ We sought to develop and validate PLM and LMM approaches for predicting functional recovery following TKA (i.e., TUG) and to compare their predictive performance. We hypothesized that PLM predictions would demonstrate superior accuracy and precision compared to LMM predictions.

## MATERIALS AND METHODS

### Description of data sources and preparation

We used two sources of post-TKA recovery data for this analysis: (1) a training dataset collected via multiple longitudinal TKA rehabilitation studies in Denver, Colorado, USA and (2) a testing dataset collected in routine practice at two physical therapy clinics in Greenville, SC, USA. Patient records in the training dataset were collected in the context of research studies and contained surgical dates from 2006 – 2017.^7,19-21^ Patient records in the testing dataset were collected in routine practice at the participating clinics and contained surgical dates from 2013 – 2018. Clinicians collected data throughout the episode of care for all patients with TKA (i.e., no selection criteria) and data were stored in a clinical database. Because the testing data were collected in real-world practice, the frequency, duration, and consistency of data collection differed across patient records. We intentionally designated our training and testing datasets to examine the robustness of PLM and LMM prediction approaches when applied to a new population with distinct geography, dates of surgery, and methods of data collection (i.e., research vs. clinical). All patient records were de-identified before being accessed for this study, and all study procedures were considered non-human subjects research (COMIRB # 18-1246).

We used the Timed Up and Go (TUG) test as the outcome of interest for our prediction approaches. The TUG is a widely used measure of post-TKA function due to its validity, responsiveness, and clinical feasibility.^22,23^ It measures the time in seconds required to complete the following steps as quickly as possible: (1) rise from a seated position, (2) walk three meters, (3) return to the chair, and (4) sit back down.^24^ Lower TUG values are associated with better patient function.

We had access to modelling variables in both datasets that are associated with post-TKA functional recovery including time since surgery, age, sex, and Body Mass Index (BMI).^10,25,26^ We included all postoperative patient records in our analyses with complete age, sex, and BMI information. We excluded records from individuals < 40 years old, observations missing TUG or time values, and observations that contained physiologically implausible values (e.g., TUG < 3 seconds). We chose to exclude observations collected > 425 days after surgery due to the sparseness of available data at later time points. See Figure 1 (Results Section) for more information related to data selection criteria.

**Figure 1.**
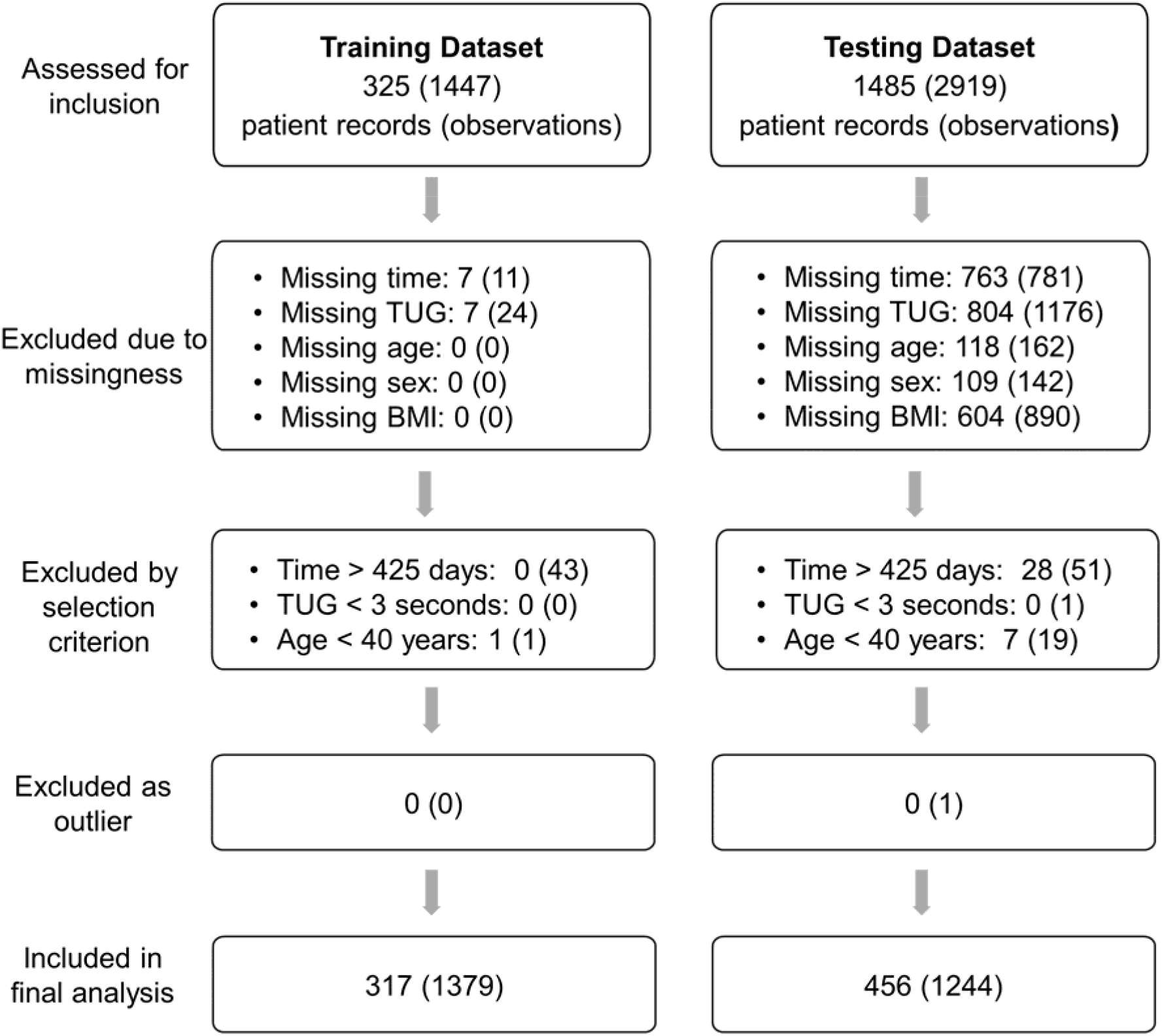
Flow diagram for assessed and included data in the final analyses

### Overview of analytical approach

We used the following steps to train and test both PLM and LMM prediction approaches: (1) build the approach using the training data, (2) examine prediction performance and tune the approach using the training data (i.e., within-sample testing), and (3) test the accuracy and precision of each approach using the testing data (i.e., out-of-sample testing). We compared performance of PLM and LMM predictions in terms of accuracy and precision across all individuals and all timepoints. We examined accuracy via the metrics of bias and coverage (Table 1). We examined precision by calculating the average width of the 50% and 80% predictions intervals. Additionally, we used mean squared error (MSE) as a summary metric of prediction performance. We calculated 95% confident intervals for each of these metrics by multiplying the standard error of the metric (across all observations) by the critical value of 1.96. Finally, we used calibration plots to visually examine predicted vs. observed TUG values in the testing data. See Table 1 for definitions of our primary accuracy and precision metrics. We used R statistical software for all analysis procedures (https://www.R-project.org/).^27^

**Table 1.**
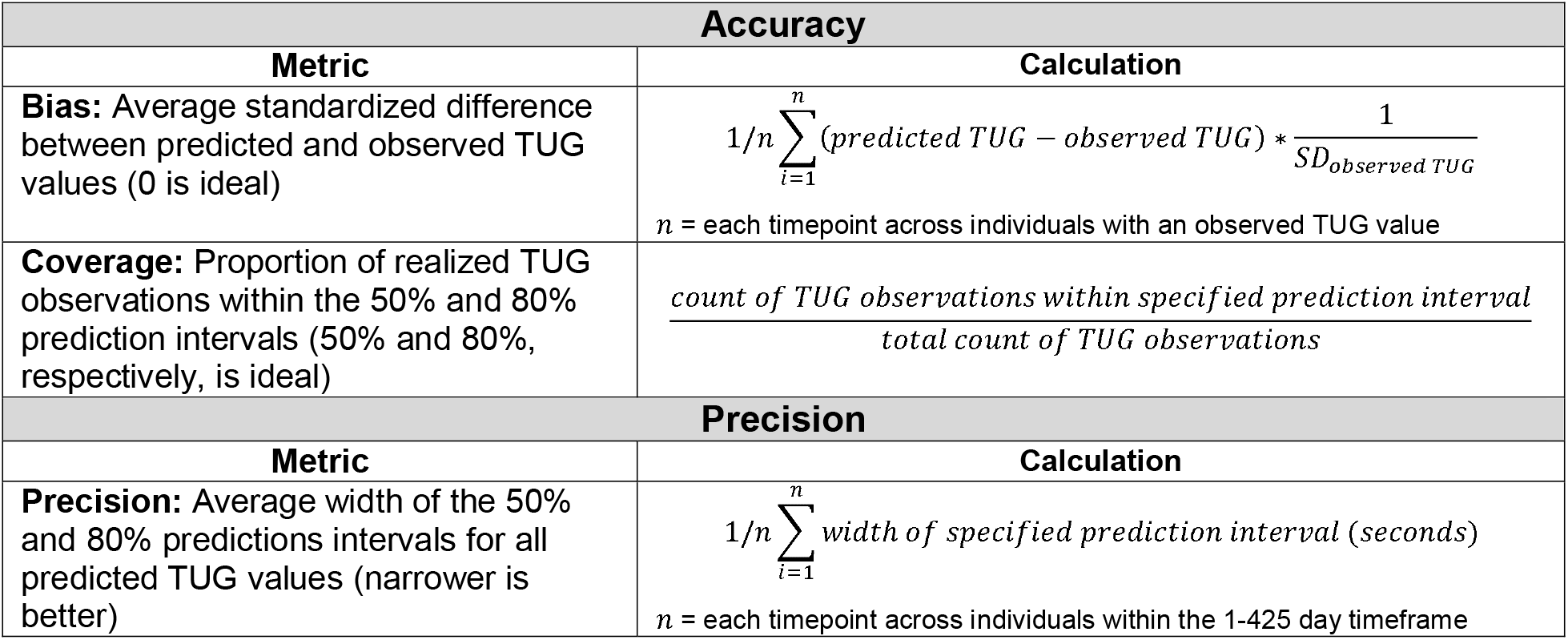
Definitions and descriptions of accuracy and precision metrics

### Training the PLM approach

#### Selecting matches using predictive mean matching

We have previously described the PLM approach in detail,^12^ but we will describe it briefly here. PLM predictions for a new patient were generated using the observed recovery data of similar historical patients. We used age, sex, BMI, baseline postoperative TUG, and baseline assessment time (days following surgery) to match an index patient to similar patients in the historical training dataset. We applied a form of predictive mean matching to determine the relative weights for each matching variable using the following steps.^28^ First, we imputed a 365-day TUG value for each patient in the training dataset via the brokenstick package in R (because data were collected at irregular timepoints).^29^ Next, we created a linear model to estimate the imputed 365-day TUG value using our matching variables of interest (see below).

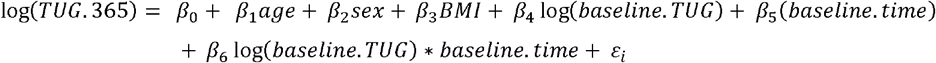

We then used this linear model to estimate the 365-day TUG value for (a) each patient in the training dataset and (b) the index patient. Finally, the patients from the training dataset with the closest predicted 365-day TUG value to the index patient were selected as matches; we used 35 patient matches based on our previous work.^12^

#### Flexibly modelling observed data from patient matches

We modeled the observed TUG data from the matching patient records to form the index patient’s predicted TUG recovery trajectory. We used Generalized Additive Models for Location, Scale, and Shape (GAMLSS) to flexibly model the median, variance, and skewness of TUG recovery from postoperative days 1-425.^30^ The GAMLSS model included a cubic spline smoother with 3 degrees of freedom for the location parameter and 1 degree of freedom for the shape parameter.

### Training the LMM approach

#### Model development

We used all available training data to build a linear mixed model for TUG recovery following TKA. First, we specified a preliminary structure for the model’s fixed effects (i.e., age, sex, BMI, postoperative time) based upon the available data and previous literature related to TKA prognosis.^10,25,26^ We adjusted for significant interaction terms amongst covariates, but we did not adjust for baseline postoperative TUG because this value was required for LMM prediction (see *LMM Prediction* section below). To capture the non-linear trajectory of TUG recovery over time, we tested plausible polynomial and spline terms including both linear and cubic splines. We considered transformations to mitigate skewness in the TUG data and examined goodness of fit. Finally, we tested plausible (a) random effects structures (i.e., random intercepts and slopes) to account for between-subjects variability and (b) within-subject correlation structures. We used Akaike Information Criterion (AIC) and likelihood ratio tests as indicated for all model specification and comparison procedures.

#### Final LMM Model

The final full LMM included all the candidate predictor variables to describe the trajectory of TUG recovery over time. We used a log transformation on the outcome to account for the skewness present in the TUG data and modeled the effect of time (standardized) using linear splines with knots placed at postoperative days 14 and 60. We specified random intercepts and slopes to account for between-subject variability and within-subject correlation. The final model is displayed below.

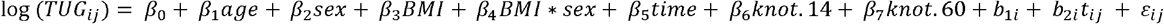

*knot.14 and knot.60 terms indicate the linear slope of TUG over time changes at postoperative days 14 and 60

\**b*_1*i*_ (intercept) and *b*_2*i*_*t*_*ij*_ (slope) indicate random effect terms

### LMM predictions

We used the IndvPred_lme function in the JMbayes package to create predictions of postoperative TUG recovery from the final LMM.^17,18,31^ The index patient’s baseline TUG observation was used to estimate the index patient’s random effect values via Monte Carlo simulation. Subsequently, the LMM was used to predict TUG values from postoperative day 1-425 for each patient. We also used the IndvPred_lme function to calculate the 50% and 80% prediction intervals for each patient at every timepoint. Additional information related to the model parameters, model summary, and functions used for the LMM prediction approach can be found in Supplementary File 1.

## RESULTS

After imposing our selection criteria, we included 317 patient records with 1379 observations in the training dataset and 456 patient records with 1244 observations in the testing dataset (Figure 1). We subsequently removed one outlier observation from the testing dataset for our analysis because it fell over 22 standard deviations away from the mean value of observations recorded in the same quartile of time. Patient demographics and dataset characteristics differed significantly in every category between training and testing datasets except for sex distribution. Patients from the training dataset were younger and demonstrated lower BMI than patients in the testing dataset. Patient records from the training dataset had more observations, longer follow up times, earlier baseline assessments, and higher baseline TUG values than patient records in the testing dataset (Table 2).

**Table 2.** Patient demographics and characteristics of training and testing datasets.

| Comparator | Training Dataset | Testing Dataset | p-value |
| --- | --- | --- | --- |
|  | 317 patient records<br>1379 observations | 456 patient records<br>1243 observations |  |
| Observations per record | 4.3 (1.4) | 2.7 (2.0) | p < 0.001* |
| Duration of assessment period, Days | 284.5 (136.6) | 122.9 (113.1) | p < 0.001* |
| Age, Years | 64.1 (7.9) | 66.4 (8.3) | p < 0.001* |
| Sex, % Female | 54% | 59% | p = 0.27 |
| BMI, kg/m<sup>2</sup> | 30.2 (5.0) | 32.7 (6.7) | p < 0.001* |
| Baseline assessment time, Days following surgery | 19.2 (13.3) | 55.3 (81.9) | p < 0.001* |
| Baseline TUG, Seconds | 28.4 (44.0) | 13.1 (7.0) | p < 0.001* |

Values presented as mean (standard deviation) unless noted otherwise.
\*Significant difference between groups via independent t-tests (continuous) or $\chi^2$ tests (categorical)

**Table 3.** Within-sample comparison of MSE, accuracy, and precision

| Metric | PLM | LMM |
| --- | --- | --- |
| MSE | 4.62 (2.46, 6.79) | 7.74 (4.93, 10.5) |
| **Accuracy** |  |  |
| bias | -0.08 (-0.12, -0.03) | <0.01 (-0.05, 0.06) |
| coverage: 50% interval | 0.51 (0.48, 0.54)* | 0.62 (0.59, 0.65) |
| coverage: 80% interval | 0.80 (0.77, 0.82)* | 0.86 (0.84, 0.88) |
| **Precision** |  |  |
| precision: 50% interval | 2.01 (1.94, 2.08)* | 2.99 (2.89, 3.10) |
| precision: 80% interval | 3.90 (3.77, 4.03)* | 5.75 (5.56, 5.95) |

Values in parentheses indicate 95% confidence intervals
\*Indicates PLM and LMM confidence interval do not overlap

**Table 4.** Out-of-sample comparison of MSE, accuracy, and precision

| Metric | PLM | LMM |
| --- | --- | --- |
| MSE | 7.42 (4.8, 10.05) | 6.30 (4.72, 7.89) |
| **Accuracy** |  |  |
| bias | $< 0.01$ (-0.05, 0.06) | 0.09 (0.04, 0.14) |
| coverage: 50% interval | 0.51 (0.47, 0.54) | 0.57 (0.54, 0.61) |
| coverage: 80% interval | 0.79 (0.76, 0.82) | 0.82 (0.80, 0.85) |
| **Precision** |  |  |
| precision: 50% interval | 2.31 (2.23, 2.39)* | 3.62 (3.53, 3.70) |
| precision: 80% interval | 4.46 (4.31, 4.61)* | 6.70 (6.84, 7.16) |

Values in parentheses indicate 95% confidence intervals
\*Indicates PLM and LMM confidence interval do not overlap

### Within-sample testing: PLM approach demonstrates superior coverage and precision

PLM and LMM approaches were similar in terms of MSE and bias via within-sample testing. Both approaches demonstrated an average bias of < 0.1 standard deviations. The PLM approach demonstrated superior coverage; the target coverage fell within the 95% confidence interval of the realized proportions for both the 50% and 80% prediction intervals. Conversely, the LMM approach demonstrated over-coverage for both 50% and 80% prediction intervals. PLM predictions were also more precise (∼33%) on average compared to LMM predictions according to both the 50% and 80% prediction intervals.

### Out-of-sample testing: PLM approach demonstrates superior coverage and precision

PLM and LMM approaches were also similar in terms of MSE and bias via out-of-sample testing. Both approaches demonstrated an average bias of < 0.1 standard deviations. The PLM approach demonstrated superior coverage; the target coverage fell within the 95% confidence interval of the realized proportions for both the 50% and 80% prediction intervals. The LMM approach also demonstrated good coverage for the 80% prediction interval but demonstrated over-coverage for the 50% prediction interval. PLM predictions were again more precise (∼34%) on average than LMM predictions.

### Prediction calibration and visualization

We examined PLM and LMM calibration across deciles of predicted TUG values. For within-sample PLM predictions, the mean predicted value fell within the confidence interval of the mean observed value across all deciles (Figure 2, Panel A). For out-of-sample PLM predictions, the mean predicted value fell within the confidence interval of the mean observed value for all deciles except the 30^th^-50^th^ deciles (Figure 2, Panel C). For within-sample LMM predictions, the mean predicted value for the highest and lowest predicted deciles fell well outside of the confidence interval for the mean observed value (Figure 2, Panel B). Out-of-sample LMM predictions followed a similar pattern, but the predicted value for five of the deciles fell outside of confidence interval for the mean observed value (Figure 2, Panel D).

**Figure 2.**
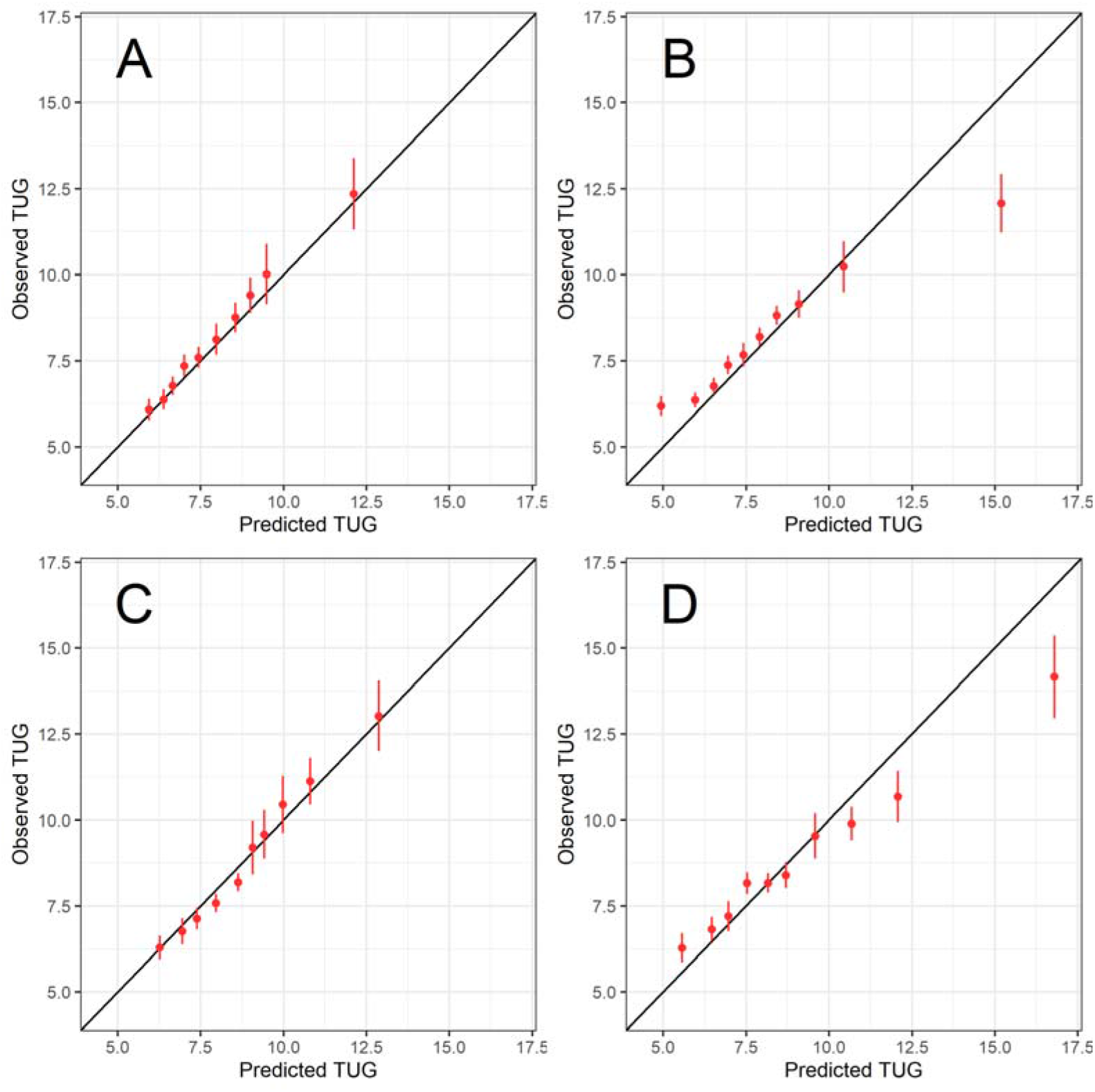
Calibration plots for PLM and LMM models in both the training and testing datasets. PLM calibration is displayed in the left column for the training dataset (Panel A) and the testing dataset (Panel C). LMM calibration is displayed in the right column for the training dataset (Panel B) and the testing dataset (Panel D). Observations were binned into deciles according to predicted TUG value. The mean observed TUG value (y axis) is plotted against the mean predicted TUG value (x axis) for each decile. The error bars indicate the confidence interval around the mean observed TUG value.

To visually probe the difference in precision between approaches, we divided the testing dataset into tertiles according to the difference between PLM and LMM precision (i.e., width of 50% interval for PLM prediction – width of 50% interval for LMM prediction). We randomly selected two patient records from each tertile and plotted the patient’s predicted TUG trajectory using both PLM and LMM approaches (Figure 3). The patient demographics and dataset characteristics of each tertile are available in Supplementary Table 1.

**Figure 3.**
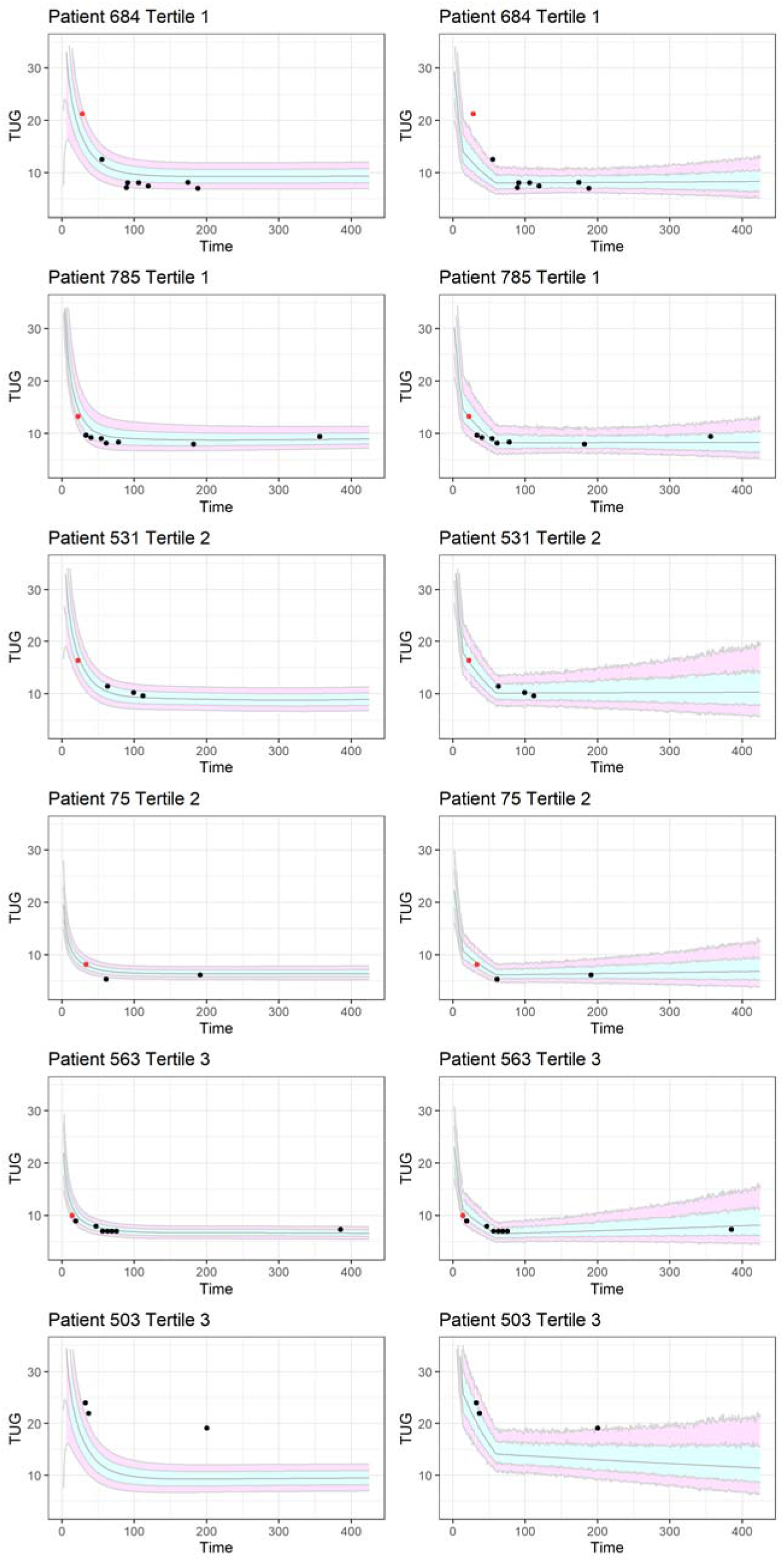
Plotting PLM (left) vs. LMM (right) prediction trajectories for individual patients PLM trajectories (left) are compared to LMM trajectories (right) for individual patient records in the testing dataset. Patient records were divided into tertiles according to the difference in PLM vs. LMM precision (i.e., PLM 50% prediction interval – LMM 50% prediction interval). Two records were randomly selected from each tertile for plotting. The black line indicates the mean predicted value and the prediction intervals are indicated by the surrounding shaded region (light blue = 50% prediction interval, lavender = 80% prediction interval). The patient’s observed TUG values are indicated by the dots. Red dots indicate the patient’s baseline TUG observation and black dots indicate non-baseline observations. Patient 503’s baseline observation is not depicted because it fell above the y-axis scale.

## DISCUSSION

This study aimed to compare PLM and LMM approaches for predicting functional recovery following TKA. Although the two approaches performed similarly in terms of MSE and average bias, the PLM approach demonstrated better overall accuracy and precision. Specifically, the PLM approach appeared well-calibrated across most deciles of predicted TUG values, while the LMM approach appeared to consistently underestimate the lowest and overestimate the highest deciles of predicted TUG values. This supports our hypothesis that PLM predictions—being constrained to previously observed values—may better accommodate the idiosyncrasies of real-world data (e.g., ceiling and floor effects). Additionally, the PLM approach consistently demonstrated accurate coverage, whereas the LMM approach demonstrated over-coverage via both within- and out-of-sample testing. Finally, the PLM approach demonstrated superior precision with prediction intervals approximately 33% more narrow than LMM prediction intervals on average. The clinical importance of the differences in PLM and LMM performance is a topic worthy of discussion and further investigation.

We designed this study with the clinical application of the predictions in mind.^32^ We envisioned clinicians could use PLM or LMM predictions to establish prognosis, help set patient expectations for functional recovery following TKA,^33,34^ and prospectively monitor patient recovery against an expected recovery trajectory.^13^ To facilitate this clinical application, we only included predictor variables that could be easily obtained in clinical practice^35^ and used modelling strategies that allowed for TUG prediction at any postoperative timepoint, so predictions can be generated whenever a patient happens to be evaluated. We also chose performance metrics to illustrate the strengths and weaknesses of PLM and LMM predictions with respect to their potential clinical application. We felt the ideal prediction approach should provide (1) an accurate, unbiased point estimate across a range of possible TUG values (i.e., bias, calibration) and (2) an accurate and precise estimate of the uncertainty around that point estimate (i.e., coverage, precision).

The importance of accurately and precisely estimating uncertainty may be an overlooked aspect of prediction in health care. Uncertainty is unavoidable in clinical practice; it should be expected that patients will rarely demonstrate exactly the point estimate of a continuous variable such as postoperative TUG. Instead, it is the degree of the deviation from what is predicted that most informs clinical decisions. What constitutes an acceptable vs. an alarming deviation requires an accurate prediction interval (i.e., coverage), not just an accurate point estimate. The PLM approach (unlike LMM) displayed nearly ideal coverage across all examined intervals, which can provide the prediction user with confidence when interpreting differences between a patient’s predicted and observed TUG values following TKA.

The precision of a prediction may also be particularly important when the outcome is interpreted on an individual basis instead of a population benchmark. Apart from population-level cut-offs for fall risk (with debatable validity in the TKA population),^36^ there are no established thresholds for “successful” TUG recovery after TKA. As with many elective procedures, this definition should be anchored to the individual’s prognosis and their postoperative goals. Therefore, it may be most useful to compare a patient’s observed TUG value to a personalized prediction for TUG recovery at a given timepoint following surgery. If the patient’s prediction interval is narrow (increased precision), the clinician can more easily detect an unexpected deviation in TUG recovery, which can then inform clinical decisions such as adjustments to a rehabilitation program. On the other hand, an inaccurately wide prediction interval may mask clinically relevant deviations in TUG recovery. The LMM approach yielded wider prediction intervals than the PLM approach and also demonstrated over-coverage. These results suggest the LMM approach yielded more uncertainty than is realistic. Conversely, the PLM approach yielded more realistic estimates of prediction uncertainty, which likely was facilitated by its semi-parametric use of previously observed data.

The statistical performance of a prediction approach is only one aspect of its clinical utility. To be clinically useful, predictions must be believable and interpretable; clinicians are less likely to trust or accept overly complex prediction models with uninterpretable parameters.^37,38^ The PLM approach may have advantages in this regard, as predictions are created from (and can be displayed alongside) the observed recovery data of previous similar patients. The LMM approach may be less interpretable to individuals unfamiliar with mixed effects regression modelling. Predictions must also be generalizable to populations outside of the development sample to be clinically useful. We found that both PLM and LMM predictions performed reasonably well in the testing dataset, which differed from the training dataset in terms of patient demographics and geography, as well as the method, cadence, and dates of data collection (Table 2).^39^ However, the PLM approach’s more consistent calibration in the testing dataset suggests it may be better equipped to handle the unique characteristics of different populations.

Prediction approaches must also be readily accessible to organizations for clinical deployment. The LMM approach may offer more accessibility than the PLM approach. The PLM approach requires access to a suitable donor dataset; creating, sharing, and maintaining these PLM datasets can be resource intensive and may pose logistical challenges. The PLM approach may also be more challenging to recalibrate as new data accrue over time because multiple, interdependent steps are required to create predictions. Conversely, LMM predictions can be created simply by using published model parameters and can be maintained using relatively straightforward model specification techniques.

Future work investigating PLM or LMM prediction approaches should evaluate (1) strategies for implementing these predictions into practice and (2) the impact of using these predictions to inform clinical decisions.^32,35^ Clinical implementation for both PLM and LMM predictions would require the development of user interfaces. Ideally, these interfaces would be integrated into the electronic medical record to complement clinicians’ typical workflow.^40-42^ The interface could use a variety of formats to support clinical decision making. The most straightforward approach would be to visually display the patient’s observed recovery after each measurement compared to their predicted recovery (e.g., Figure 3). Alternatively, the interface could alert the clinician when a patient’s observed recovery falls outside of a predefined interval (e.g., 80% prediction interval), which could prompt the clinician to consider whether a change in the treatment plan is warranted. Regardless of the chosen implementation strategy, the clinical impact of using PLM or LMM predictions should be prospectively compared to the standard of care in future work.^4,35,37^

## Limitations

Our study design has a few limitations. We used a limited number of variables to develop both prediction approaches. Including additional variables may have improved both PLM and LMM performance, but the increased burden placed on clinicians to capture these variables may also limit the feasibility of using the predictions in practice. The generalizability of our analysis may also be limited by the extent of missingness in the testing dataset (see Supplementary Table 2 for demographics of excluded patient records). This missingness may have been inflated by preoperative data that we unknowingly assessed for inclusion. We could not determine whether observations that were missing date values (i.e., date of surgery, date of assessment) were taken pre- or postoperatively. We have previously observed that preoperative observations from this database tend to contain more missingness than postoperative observations.^12^ Nonetheless, clinical databases (like the testing dataset) frequently contain missing values because data is collected to support patient care instead of research goals.^43^ However, using clinical data also promotes the inclusion of understudied patients that are often excluded from research studies.^25^ The testing dataset also provided a methodologically and geographically distinct sample for examining PLM and LMM prediction performance, which we believe strengthens our findings. Regardless, future research should include prospective assessment of PLM and LMM prediction performance.

## Conclusion

Overall, the PLM approach provided more accurate and precise estimates of post-TKA functional recovery compared to the LMM approach. These results suggest that PLM predictions may be more useful and generalizable in post-TKA clinical care. However, PLM and LMM approaches differ in characteristics that may affect their ability to be deployed clinically. Before pursuing a prediction approach, organizations should consider the approach’s intended use, interpretability, resources required for deployment, and fit within the clinical workflow. Future work involving PLM or LMM predictions of post-TKA recovery should (1) prospectively assess prediction performance, (2) develop and test user interfaces to translate predictions into clinical care, and (3) evaluate the impact of using these predictions on health outcomes.^37,44^

## Data Availability

The datasets used for this analysis may be available upon reasonable request. Please contact the senior author, Dr. Jennifer Stevens-Lapsley, for details.

