## Supplemental File 1 for "Comparing “people-like-me” and linear mixed model predictions of functional recovery following knee arthroplasty"

### Supplemental Materials: Linear Mixed Model Summary and Prediction

#### Required packages

```
library(tidyverse)
library(magrittr)
library(nlme)
library(JMbayes)
library(lspline)
library(knitr)
library(kableExtra)
```

#### Final Linear Mixed Model

```
#read in data
dat <- readRDS("Data/training_dat_final.rds")


# Standardize time and format id as factor
dat %<>% mutate(id = as.factor(id),
                time.stand = as.numeric((time - mean(time))/sd(time)))


# Final LMM model
lmm.final <- lme(log(tug) ~ 1 + bmi*sex + age + lspline(time.stand, knots = c(-0.8999027, -0.5233489)),
                       random = list(id = pdDiag(~1+ time.stand)),
                       data = dat,
                       method = "REML")

# Note: knot values of c(-0.8999027, -0.5233489) are the standardized values of c(14, 60)


coef(summary(lmm.final)) %>% kbl(booktabs = T)
```

| | Value | Std.Error | DF | t-value | p-value |
|---|---|---|---|---|---|
| (Intercept) | -11.8005261 | 0.3180905 | 1059 | -37.098019 | 0.0000000 |
| bmi | 0.0081242 | 0.0043244 | 312 | 1.878695 | 0.0612184 |
| sex2 | -0.2437595 | 0.1693580 | 312 | -1.439315 | 0.1510637 |
| age | 0.0117493 | 0.0017090 | 312 | 6.874799 | 0.0000000 |
| lspline(time.stand, knots = c(-0.8999027, -0.5233489))1 | -14.6932797 | 0.2754183 | 1059 | -53.348962 | 0.0000000 |
| lspline(time.stand, knots = c(-0.8999027, -0.5233489))2 | -1.1571083 | 0.0561612 | 1059 | -20.603333 | 0.0000000 |
| lspline(time.stand, knots = c(-0.8999027, -0.5233489))3 | -0.0307099 | 0.0072268 | 1059 | -4.249439 | 0.0000233 |
| bmi:sex2 | 0.0122935 | 0.0054924 | 312 | 2.238258 | 0.0259094 |

#### Create predictions from the final LMM

```
# Create vector of time from 1:425 and convert to standardized time
time.vector <- c(1:425)
time.vector <- (time.vector - mean(dat$time))/sd(dat$time)


# Create dataframe with only baseline TUG values
dat.baseline <- dat %>% group_by(id) %>% arrange(time) %>% slice(1L)


# Create predictions using baseline TUG value with 50% prediction interval

m.pred.5 <- IndvPred_lme(lmeObject = lmm.final,
              newdata = dat.baseline,
              timeVar = c("time.stand"),
              all_times = TRUE,
              return_data = TRUE,
              interval = "prediction",
              times = time.vector,
              level = 0.5,
              M = 500)


# Create predictions using baseline TUG value with 80% prediction interval

m.pred.9 <- IndvPred_lme(lmeObject = lmm.final,
                          newdata = dat.baseline,
                          timeVar = "time.stand",
                          all_times = TRUE,
                          return_data = TRUE,
                          interval = "prediction",
                          times = time.vector,
                          level = 0.8,
                          M = 500)


# convert predictions out of log scale
m.pred.5 %<>% mutate(C50 = exp(pred),
                     C75 = exp(upp),
```

```
                      C25 = exp(low))

m.pred.9 %<>% mutate(C50 = exp(pred),
                      C90 = exp(upp),
                      C10 = exp(low))
```
