## Supplementary Tables for "Comparing “people-like-me” and linear mixed model predictions of functional recovery following knee arthroplasty"

**Supplementary Table 1.** Demographics and characteristics of the testing dataset divided into tertiles according to the difference between PLM and LMM precision.*

| **Comparator** | **1^st^ tertile** | **2^nd^ tertile** | **3^rd^ tertile** | **p-value** |
| --- | --- | --- | --- | --- |
| Difference in 50% precision interval (PLM – LMM) | -0.6 | -1.3 | -1.8 | <0.001 |
| Difference in 80% precision interval (PLM – LMM) | -1.1 | -2.5 | -3.5 | <0.001 |
| Age, Years | 66.8 | 66.8 | 65.5 | 0.25 |
| Sex, % Female | 51.3% | 57.2% | 67.1% | 0.025 |
| BMI, kg/m^2^ | 31.3 | 32.7 | 33.3 | <0.001 |
| Baseline assessment time, Days following surgery | 106.0 | 32.6 | 27.4 | <0.001 |
| Baseline TUG, Seconds | 10.6 | 13.3 | 15.6 | <0.001 |

*The first tertile represents patient records with the smallest difference in precision for the 50% prediction interval (i.e., PLM 50% prediction interval – LMM 50% prediction interval). The third tertile represents patient records with the largest difference in 50% prediction intervals between approaches.

**Supplementary Table 2.** Patient demographics for included and excluded patient records in the testing dataset.

| **Comparator** | **Included**  **testing data** | **Excluded testing data** | **p-value** |
| --- | --- | --- | --- |
|  | 456 records  1244 observations | 1206 records  1675 observations |  |
| Age, Years | 66.4 (8.3) | 65.9 (9.2) | p = 0.31 |
| Sex, % Female | 59% | 60% | p = 0.662 |
| BMI, kg/m^2^ | 32.7 (6.7) | 31.4 (7.0) | p = 0.02* |

Values presented as mean (standard deviation) unless noted otherwise.

*Indicates statistically significant difference between groups. Independent t-tests were used to compare continuous variables and chi-squared tests were used for categorical variables.
